# Failure to replicate the association of rare loss-of-function variants in type I IFN immunity genes with severe COVID-19

**DOI:** 10.1101/2020.12.18.20248226

**Authors:** Gundula Povysil, Guillaume Butler-Laporte, Ning Shang, Chen Weng, Atlas Khan, Manal Alaamery, Tomoko Nakanishi, Sirui Zhou, Vincenzo Forgetta, Robert Eveleigh, Mathieu Bourgey, Naveed Aziz, Steven Jones, Bartha Knoppers, Stephen Scherer, Lisa Strug, Pierre Lepage, Jiannis Ragoussis, Guillaume Bourque, Jahad Alghamdi, Nora Aljawini, Nour Albes, Hani M. Al-Afghani, Bader Alghamdi, Mansour Almutair, Ebrahim Sabri Mahmoud, Leen Abu Safie, Hadeel El Bardisy, Fawz S. Al Harthi, Abdulraheem Alshareef, Bandar Ali Suliman, Saleh Alqahtani, Abdulaziz AlMalik, May M. Alrashed, Salam Massadeh, Vincent Mooser, Mark Lathrop, Yaseen Arabi, Hamdi Mbarek, Chadi Saad, Wadha Al-Muftah, Radja Badji, Asma Al Thani, Said I. Ismail, Ali G. Gharavi, Malak S. Abedalthagafi, J Brent Richards, David B. Goldstein, Krzysztof Kiryluk

**Author notes:** co-corresponding authors: J Brent Richards, David B. Goldstein and Krzysztof Kiryluk.

## Abstract

A recent report found that rare predicted loss-of-function (pLOF) variants across 13 candidate genes in TLR3- and IRF7-dependent type I IFN pathways explain up to 3.5% of severe COVID-19 cases. We performed whole-exome or whole-genome sequencing of 1,934 COVID-19 cases (713 with severe and 1,221 with mild disease) and 15,251 ancestry-matched population controls across four independent COVID-19 biobanks. We then tested if rare pLOF variants in these 13 genes were associated with severe COVID-19. We identified only one rare pLOF mutation across these genes amongst 713 cases with severe COVID-19 and observed no enrichment of pLOFs in severe cases compared to population controls or mild COVID-19 cases. We find no evidence of association of rare loss-of-function variants in the proposed 13 candidate genes with severe COVID-19 outcomes.

## Main Text

Host genetic factors related to COVID-19 susceptibility and outcomes are of great interest, as their identification could elucidate the mechanisms of SARS-CoV-2 infection and severity, thereby providing clues about potential therapeutic targets. Recently, Zhang *et al*.^1^ reported deleterious mutations in 13 candidate genes involved in type I IFN immunity to be associated with COVID-19 severity. The authors reported a significant enrichment in predicted loss-of-function (pLOF) variants in 659 severe COVID-19 cases relative to 534 controls with asymptomatic or benign infection using a burden test under a dominant model of inheritance (odds ratio (OR) = 8.28 [1.04-65.64], p=0.01). In addition to the nine pLOFs in cases used in the burden test, they detected 109 missense or in-frame indels and tested them experimentally in ad hoc overexpression systems; 24 variants (including all the pLOFs) were found to be hypomorphic and were detected in 23 patients of various ages (17-77 years) and of both sexes. The authors reported the presence of one pLOF in controls but did not provide the total number of other functional variants, or whether variants identified in controls were experimentally tested as well. The authors concluded that rare deleterious variants in the TLR3, IRF7 and IRF9-dependent type 1 interferon pathway genes explain up to 3.5% of severe cases of COVID-19.

We aimed to replicate these findings using independent datasets. We note that an exact replication of all the reported associations is not possible because the study design of Zhang et al does not permit a formal statistical comparison of the same variant types in cases and controls. The reason for this is that missense variants were reported as functionally characterized in cases but not controls, so the counts of functionally compromised missense variants cannot be evaluated in a statistical test. In order to test the role for rare variation in these genes in COVID-19 outcomes, we therefore performed association tests on all rare variation that is apparently functional, and specifically focused on predicted loss-of-function variants, applying identical rules to case and control variants, as is appropriate in such analyses.

Our first analysis was based on the cohort of patients recruited by the Columbia University COVID-19 Biobank (**Tables 1 and 2**). We performed exome sequencing of 1,153 COVID-19 cases of diverse ethnicities who were treated at Columbia University Irving Medical Center at the peak of the New York pandemic, between March and May, 2020. Of 1,153 cases, 480 had severe COVID-19 that led to death or required endotracheal intubation due to respiratory failure (**Table 2** and **Methods**). In an attempt to replicate the findings by Zheng et al., we performed a gene-set based collapsing association test stratified by ancestry using 9,589 population controls of similar ancestries (see **Methods**).

**Table 1.** Study Cohorts

|  | **COVID-19 Cases** | **Severe COVID-19** | **Mild COVID-19** | **Population Controls** |
| --- | --- | --- | --- | --- |
| Columbia | 1,153 | 480 | 673 | 9,589 |
| Quebec | 220 | 62 | 158 | 313 |
| Saudi Arabia | 307 | 148 | 159 | 218 |
| Qatar | 254 | 23 | 231 | 5,131 |
| Total | 1,934 | 713 | 1,221 | 15,251 |

**Table 2.** Cohort characteristics.

|  | Columbia University<br>COVID-19 Biobank |  | Biobanque Québec<br>COVID-19 |  | Saudi Arabia<br>COVID-19 Biobank |  | Qatar Genome Program<br>COVID-19 |  |
| --- | --- | --- | --- | --- | --- | --- | --- | --- |
|  | Severe Cases<br>N=480 | Mild Cases<br>N=673 | Severe Cases<br>N=62 | Mild Cases<br>N=158 | Severe Cases<br>N=148 | Mild Cases<br>N=159 | Severe Cases<br>N=23 | Mild Cases<br>N=231 |
| **Age** |  |  |  |  |  |  |  |  |
| 0-9 | 3 (0.62%) | 9 (1.34%) | 0 (0.00%) | 0 (0.00%) | 1 (0.68%) | 4 (2.52%) | 0 (0.00%) | 0 (0.00%) |
| 10-19 | 9 (1.88%) | 5 (0.74%) | 0 (0.00%) | 0 (0.00%) | 1 (0.68%) | 8 (5.03%) | 0 (0.00%) | 2 (0.87%) |
| 20-29 | 5 (1.04%) | 34 (5.05%) | 2 (3.23%) | 6 (3.80%) | 0 (0.00%) | 56 (35.22%) | 1 (4.35%) | 57 (24.68%) |
| 30-39 | 16 (3.33%) | 66 (9.81%) | 2 (3.23%) | 10 (6.33%) | 10 (6.76%) | 36 (22.64%) | 3 (13.04%) | 75 (32.47%) |
| 40-49 | 28 (5.83%) | 60 (8.92%) | 2 (3.23%) | 13 (8.23%) | 12 (8.11%) | 14 (8.81%) | 7 (30.43%) | 45 (19.48%) |
| 50-59 | 62 (12.92%) | 124 (18.42%) | 9 (14.52%) | 27 (17.09%) | 20 (13.51%) | 22 (13.84%) | 7 (30.43%) | 37 (16.02%) |
| 60-69 | 97 (20.21%) | 137 (20.36%) | 13 (20.97%) | 20 (12.66%) | 51 (34.46%) | 8 (5.03%) | 5 (21.74%) | 11 (4.76%) |
| 70-79 | 133 (27.71%) | 130 (19.32%) | 17 (27.42%) | 24 (15.19%) | 34 (22.97%) | 9 (5.66%) | 0 (0.00%) | 4 (1.73%) |
| 80-89 | 89 (18.54%) | 72 (10.70%) | 11 (17.74%) | 40 (25.32%) | 17 (11.49%) | 2 (1.26%) | 0 (0.00%) | 0 (0.00%) |
| 90-99 | 30 (6.25%) | 23 (3.42%) | 5 (8.06%) | 15 (9.49%) | 2 (1.35%) | 0 (0.00%) | 0 (0.00%) | 0 (0.00%) |
| ≥ 100 | 1 (0.21%) | 0 (0.00%) | 0 (0.00%) | 0 (0.00%) | 0 (0.00%) | 0 (0.00%) | 0 (0.00%) | 0 (0.00%) |
| Unknown | 7 (1.46%) | 13 (1.93%) | 1 (1.61%) | 3 (1.90%) | 0 (0.00%) | 0 (0.00%) | 0 (0.00%) | 0 (0.00%) |
| **Gender** |  |  |  |  |  |  |  |  |
| Male | 288 (60.00%) | 357 (53.05%) | 44 (70.97%) | 64 (40.51%) | 100 (67.57%) | 89 (55.97%) | 13 (56.52%) | 116 (50.22%) |
| Female | 192 (40.00%) | 316 (46.95%) | 18 (29.03%) | 94 (59.49%) | 48 (32.43%) | 70 (44.03%) | 10 (43.48%) | 115 (49.78%) |
| **Ancestry** |  |  |  |  |  |  |  |  |
| African | 192 (40.00%) | 272 (40.42%) | 9 (14.52%) | 21 (13.29%) | 0 (0.00%) | 0 (0.00%) | 0 (0.00%) | 0 (0.00%) |
| East Asian | 10 (2.08%) | 16 (2.38%) | 6 (9.68%) | 16 (10.13%) | 0 (0.00%) | 0 (0.00%) | 0 (0.00%) | 0 (0.00%) |
| European | 27 (5.62%) | 40 (5.94%) | 45 (72.58%) | 109 (68.99%) | 0 (0.00%) | 0 (0.00%) | 0 (0.00%) | 0 (0.00%) |
| Latino | 217 (45.21%) | 289 (42.94%) | 2 (3.23%) | 5 (3.16%) | 0 (0.00%) | 0 (0.00%) | 0 (0.00%) | 0 (0.00%) |
| Middle Eastern | 30 (6.25%) | 52 (7.73%) | 0 (0.00%) | 0 (0.00%) | 148 (100.00%) | 159 (100.00%) | 23 (100.00%) | 231 (100.00%) |
| South Asian | 3 (0.62%) | 4 (0.59%) | 0 (0.00%) | 7 (4.43%) | 0 (0.00%) | 0 (0.00%) | 0 (0.00%) | 0 (0.00%) |
| Admixed | 1 (0.21%) | 0 (0.00%) | 0 (0.00%) | 0 (0.00%) | 0 (0.00%) | 0 (0.00%) | 0 (0.00%) | 0 (0.00%) |
| **Comorbidities\*** |  |  |  |  |  |  |  |  |
| Diabetes | 194 (40.42%) | 214 (31.80%) | 25 (40.32%) | 45 (28.48%) | 91 (61.49%) | 18 (11.32%) | 9 (39.13%) | unknown |
| Chronic Kidney Disease | 111 (23.13%) | 109 (16.20%) | 11 (17.74%) | 20 (12.66%) | 19 (12.84%) | 1 (0.63%) | 2 (8.70%) | unknown |
| Chronic Lung Disease | 139 (28.96%) | 142 (21.10%) | 14 (22.58%) | 25 (15.82%) | 18 (12.16%) | 6 (3.77%) | 2 (8.70%) | unknown |
| Chronic Heart Disease | 132 (27.50%) | 152 (22.59%) | 14 (22.58%) | 19 (12.03%) | unknown | unknown | 1 (4.35%) | unknown |
| Cancer | 131 (27.29%) | 174 (25.85%) | 5 (8.06%) | 7 (4.43%) | 5 (3.38%) | 0 (0.00%) | 0 (0.00%) | unknown |

\* Note: Chronic Lung Disease includes asthma, chronic obstructive pulmonary disease, interstitial pulmonary disease, primary pulmonary hypertension; Chronic Heart Disease includes coronary artery disease and heart failure.

When testing for pLOF variants with an internal and external minor allele frequency of less than 0.001 in any of the 13 genes of interest (see **Methods**), we did not detect any variant enrichment in cases (OR = 1.10 [0.03-7.67], p = 1). In fact, only one of the cases and 23 of the controls had a qualifying pLOF variant (0.21% vs. 0.24%). We also note that the carrier frequency in controls was dependent on genetic ancestry and ranged from 0 to 0.43% across different ancestral clusters. Interestingly, one of the pLOFs reported by Zhang et al. to be present in their cases was present in multiple controls from the Columbia University COVID-19 Biobank (**Table 3**). The addition of rare missense variants in the functional model also did not improve enrichment (OR = 1.22 [0.84-1.71], p = 0.3).

**Table 3.**
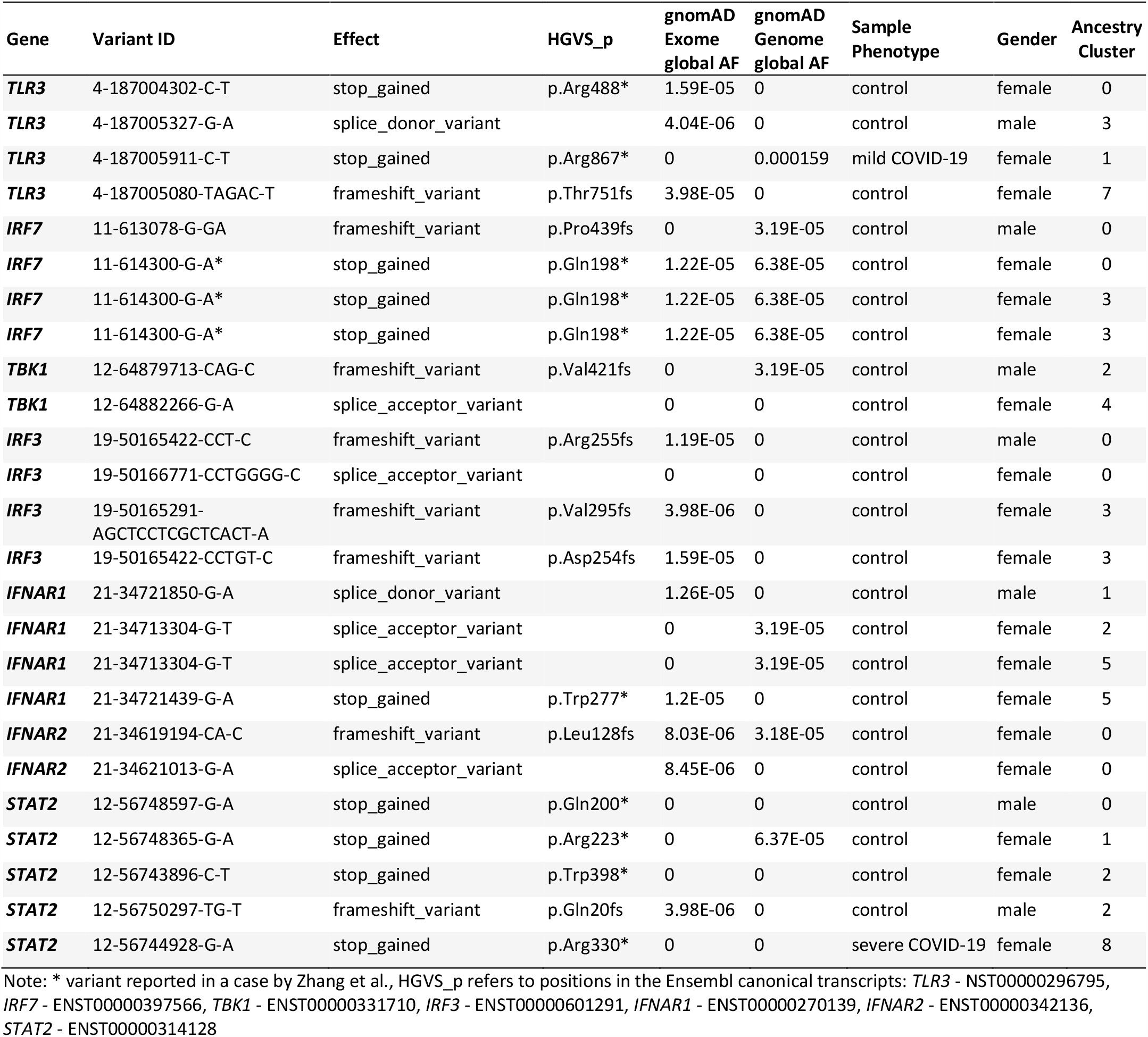
The complete list of all qualifying pLOF variants found in 1,153 COVID-19 cases (673 mild and 480 severe) and 9,589 controls from the Columbia COVID-19 Biobank cohort along with the observed allelic frequencies.

We additionally tested 480 patients with severe disease against 673 patients with milder disease that recovered from COVID-19 without the need for intubation, and again we observed no clear enrichment (OR = 1.45, [0.02-113.51], p = 1). Similarly, even when adding missense variants, we did not detect an enrichment (OR = 1.13 [0.74-1.72], p = 0.6). We also looked for any variants in the 13 genes that were listed as pathogenic or likely pathogenic in ClinVar but could not find any in our 480 severe cases. Based on our power calculations, we have over 80% power to detect odds ratios of 5.5 or greater for rare pLOF variants at an alpha of 0.05. We are therefore well powered to replicate the findings by Zhang et al. given their reported odds ratio of 8.0 (**Figure 1**).

**Figure 1.**
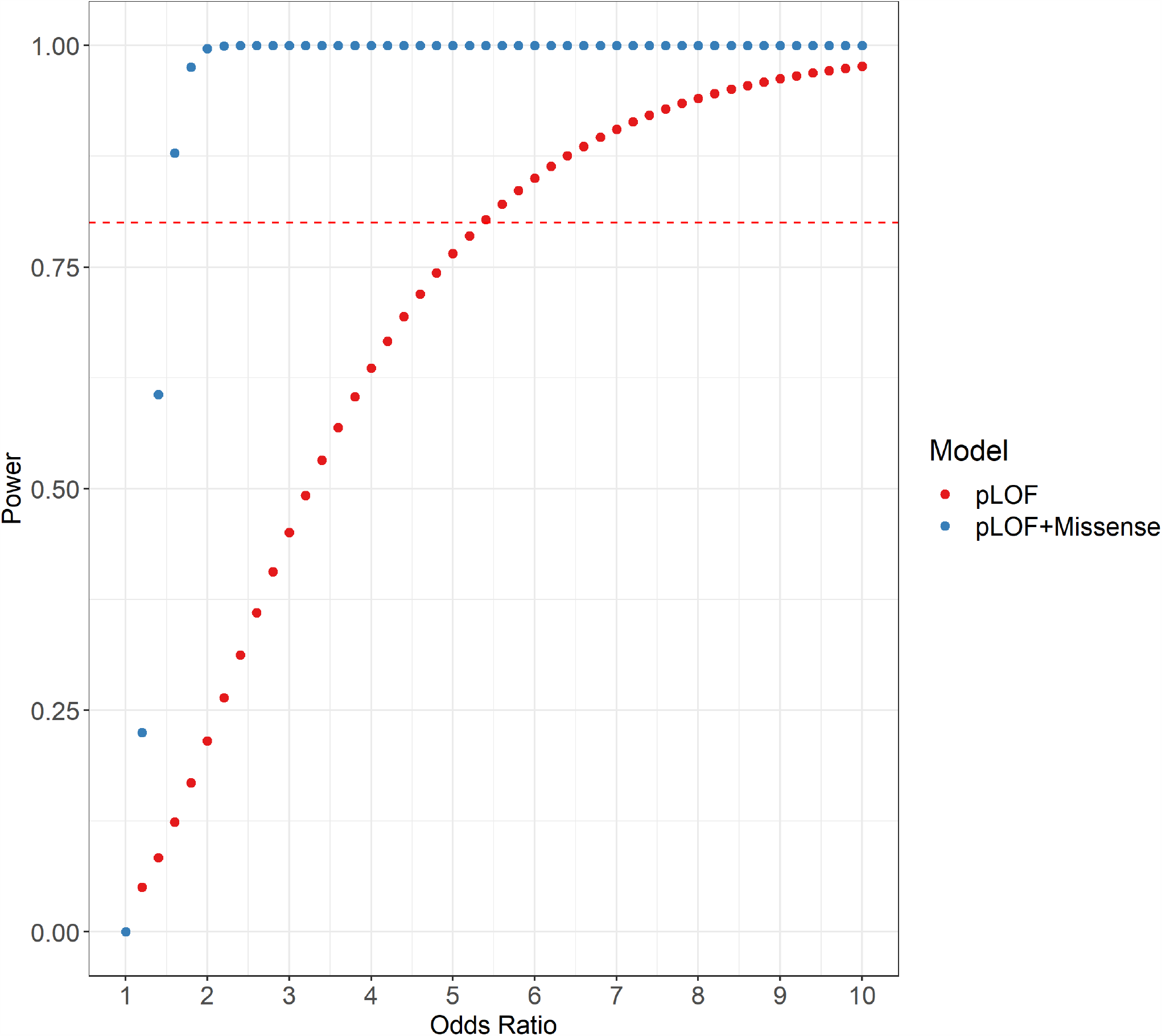
Power curves for a gene set based collapsing test: Power calculations for the Columbia COVID-19 Biobank cohort of 480 severe COVID-19 cases and 9,589 population controls were performed using the samplesizeCMH R package (version 0.0.0) for a dominant model at alpha=0.05 and a range of odds ratios (OR). Results are shown for the pLOF and model including pLOFs and missense model. Since power is influenced by the carrier frequency, we have adequate power to detect effect sizes as small as 1.5 for the model including missense variants compared to 5.5 for the rare pLOF variants only model.

The second cohort included patients recruited by the Biobanque Québec COVID-19. In total, 533 participants were recruited at the Jewish General Hospital in Montreal and underwent genome sequencing in partnership with the Canadian HostSeq project. This cohort included 62 severe COVID-19 cases with respiratory failure requiring invasive ventilatory support. An additional 471 patients were treated as controls and included 158 individuals with mild COVID-19 that did not require ventilatory support and 313 SARS-CoV-2 PCR-negative participants. We tested for enrichment of pLOF variants in the 62 severe COVID-19 cases compared to the 471 controls (see **Methods**) but did not detect a single pLOF in any of the cases or controls, even when we used a more relaxed minor allele frequency threshold of less than 1%. Similarly, we observed no significant enrichment under the missense model (OR = 1.24 [0.36-3.46], p = 0.59).

The third cohort was recruited in Saudi Arabia and sequenced in partnership with the Saudi Genome project. Exome sequencing was performed in 307 patients with COVID-19, including 148 severe cases, 159 participants with mild COVID-19 and 218 COVID-19 PCR negative controls. (see **Methods**). We could not find any cases with a rare pLOF or missense variant in any of the 13 tested genes.

The fourth cohort was collected by the Qatar Genome Project. Of 14,060 Qatar biobank participants with genome sequence data, there were 700 patients with COVID-19 of which 60 were defined as severe cases and 640 were mild or asymptomatic cases (see **Methods**). Limiting the analysis to unrelated individuals, there were 5,385 participants including 23 severe COVID-19 and 231 mild or asymptomatic COVID-19 samples. We did not find any individuals with rare pLOF variants in the 13 genes analyzed and also could not find any with a rare missense variant in COVID-19 samples.

In a meta-analysis of all four cohorts, we were also not able to detect any significant enrichment for pLOFs (OR = 1.10 [0.03-7.67], p = 1) or pLOFs and missense variants combined (OR = 1.17 [0.83-1.63], p = 0.34). In summary, in our analysis of four international COVID-19 biobanks including a total of 1,934 sequenced COVID-19 cases (713 severe and 1,221 mild), we could only find a single rare pLOF variant in a severe case and another single rare pLOF variant in a case with mild disease. We therefore observed no enrichment in pLOF variants in type I IFN immunity genes among cases of severe COVID-19 compared to mildly affected patients or to ancestry-matched population controls.

These results are further corroborated by a recent analysis of exomes from participants in the UK Biobank performed by Regeneron Inc.^2^ In the association testing of 1,184 COVID-19 cases vs. 422,318 controls, there was no association between pLOFs at these genes after careful adjustment for population stratification and multiple testing. Moreover, there was no association signal when the association tests were repeated including rare deleterious missense variants, or when the case group was limited only to severe COVID-19 cases (N=471).

Between this study and the independent report by Regeneron Inc, there were a total of 3,118 cases of COVID-19, including 1,184 cases with severe disease used to test this hypothesis. Despite large sample size, our collective results did not support the key conclusion by Zheng et al. that 3.5% of severe COVID-19 cases are explained by rare inborn errors in type I interferon immunity genes. While the age of our case cohort is slightly higher (mean age 65.9 years in our study vs. mean age 51.8 years in Zheng et al), the age of variant carriers reported by Zheng et al. ranged from 17 to 77 years, and the great majority of our cases fall within this range. Further, we have not undertaken any functional studies of missense variants identified in our cohorts. We also recognize that the inheritance pattern for deleterious variants in type I IFN immunity genes is often recessive, and that the penetrance of such variants might be dependent on male sex. However, Zheng et al. found bi-allelic variants only in a small number of cases and analyzed deleterious variants in both sexes jointly. Taken together, our negative results suggest that the findings by Zheng et al are not generalizable and highlight the need to rigorously adhere to accepted study design principles when reporting new genetic associations for a set of candidate genes^3,4^.

## Methods

### Columbia University COVID-19 Biobank Cohort

The study includes a multiethnic cohort of 1,153 COVID-19 patients treated at the Columbia University Irving Medical Center. All cases had PCR-confirmed SARS-CoV-2 infection. The patients were recruited to the Columbia University COVID-19 Biobank between March and May 2020, at the peak of the New York pandemic. A total of 480 patients were classified as having severe COVID-19. All 480 severe COVID-19 cases were defined by either death from COVID-19 (N=317), or acute respiratory failure due to COVID-19 requiring endotracheal intubation (N=163). This cohort was composed of 288 males and 192 females; the average age was 67 (range 2-101); 45% of the participants were predicted to be Hispanic/Latino, 40% African, 6% Middle Eastern, 6% European, 2% East Asian, 0.6% South Asian, and 0.2 % Admixed (see below for details about ancestry determination).

For comparisons of variant frequencies, we used an internal dataset of 9,589 local population controls with exome and genome sequence data generated by Columbia University’s Institute for Genomic Medicine (IGM). The controls are derived from the same general patient population as the cases, served by the Columbia University Irving Medical Center and sequenced as controls or healthy family members for other studies. Additional tests were performed against a cohort of 673 patients with mild COVID-19 who were recruited to Columbia COVID-19 Biobank during the same time period, but who have recovered without the need for endotracheal intubation. This group was composed of 357 males and 316 females; the average age was 59 (range 3-97); 43% of the participants were predicted to be Hispanic/Latino, 40% African, 8% Middle Eastern, 6% European, 2% East Asian, and 0.6% South Asian.

All case exomes were captured with the IDT xGen Exome Research Panel V1.0 (Integrated DNA Technologies, Coralville, IA, USA) and sequenced on Illumina’s NovaSeq 6000 (Illumina, San Diego, CA, USA) platform with 150 bp paired-end reads according to standard protocols. Exome sequencing of controls were performed on Illumina’s HiSeq 2000, HiSeq 2500, and NovaSeq 6000 using various exome capture kits. Genome controls were all sequenced on Illumina’s NovaSeq 6000.

All cases and controls were processed with the same bioinformatic pipeline for variant calling. In brief, reads were aligned to human reference GRCh37 using DRAGEN (Edico Genome, San Diego, CA, USA)^5^ and duplicates were marked with Picard (Broad Institute, Boston, MA, USA). Variants were called according to the Genome Analysis Toolkit (GATK - Broad Institute, Boston, MA, USA) Best Practices recommendations v3.6^6,7^. Finally, variants were annotated with ClinEff^8^ and the IGM’s in-house ATAV^9^ (https://github.com/igm-team/atav) was used to add custom annotations including gnomAD v2.1 frequencies^10^ and clinical annotations provided by the Human Gene Mutation Database (HGMD)^11^, ClinVar^12,13^, and Online Mendelian Inheritance in Man (OMIM). A centralized database was used to store variant and per site coverage data for all samples enabling well controlled analyses without the need of generating jointly called VCF files (see Ren et al. 2020^9^ for details). For each patient, we performed ancestry classification into one of the six major ancestry groups (European, African, Latino, East Asian, South Asian and Middle Eastern) using a neural network trained on a set of 1000 Genomes samples with known ancestry labels. We used a 50% probability cut-off to assign an ancestry label to each sample and labeled samples that did not reach 50% for any of the ancestral groups as “Admixed”.

All included samples had at least 90% of the consensus coding sequence (CCDS release 20)^14^ covered at 10x or more. Samples had ≤ 3% contamination levels according to VerifyBamID^15^. Additionally, we removed samples with a discordance between self-declared and sequence-derived gender. We used KING^16^ for the detection of related individuals and removed one of each pair that had an inferred relationship of second-degree or closer while favoring the inclusion of COVID-19 cases over controls.

For ancestry adjustment, we performed Principal Component Analysis on a set of predefined variants as previously described in Cameron-Christie et al.^17^ To identify clusters that reflect ancestry we applied the Louvain method of community detection^18^ to the first 6 principal components (PCs) resulting in 10 clusters reflecting not only continental populations, but also more detailed subdivision of Europeans. We performed coverage harmonization (see below) and collapsing within the clusters so that population specific variants would be easier to filter out.

Coverage differences between cases and controls caused by different capture kits or sequencing depth in general can potentially introduce a bias in the analysis because without sufficient coverage no variants can be called^4^. Therefore, we used the site-based pruning approach described in Petrovski et al. 2015^19^ and removed sites with an absolute difference in percentages of cases compared to controls with at least 10x coverage of greater than 7.0%.

Using the genes tested by Zhang *et al*. (*TLR3, UNC93B1, TICAM1, TRAF3, TBK1, IRF3, IRF7, IFNAR1, IFNAR2, STAT1, STAT2, IRF9*, and *IKBKG*), we performed a gene set-based collapsing analysis to test for a significant difference in the proportion of cases carrying at least one qualifying variant (QV) in the gene set compared to controls. A QV was defined as a variant that passes certain filter criteria. For the pLOF model we used basic quality control filters (e.g. GATK’s Variant Quality Score Recalibration, QUAL, GQ, QD, …), an internal and external minor allele frequency (MAF) of 0.001 and restricted the variant effect to frameshift, stop gained, splice acceptor, and splice donor labeled high confidence by LOFTEE^10^. For the internal MAF filter we used a leave-one-out filter, that ensures that singletons are retained even if the sample size is small. For the external MAF filtering we used the continental MAFs provided by gnomAD^10^ and ExAC^20^. For the functional model, we also included missense variants in addition to the pLOF variants while keeping all other filters the same. From the results of the individual clusters, we extracted the number of cases/controls with and without a QV in the gene-set and used the exact two-sided Cochran-Mantel-Haenszel (CMH) test^21,22^ to test for an association between disease status and QV status while controlling for cluster membership.

For the purpose of the meta-analysis of all four cohorts, we restricted the Biobanque Québec cohort (see below) to individuals of European ancestry and only included population controls, so the final analysis included a total of 696 severe COVID-19 cases and 15,047 population controls. We used the CMH test as described above and treated the other cohorts as additional clusters, since each of the extra cohorts was composed of a single well-defined ancestry. The CMH odds ratios and 95% confidence intervals were reported.

### Biobanque Québec COVID-19 Cohort

The Biobanque Québec COVID-19 (www.BQC19.ca) is a provincial biobank that prospectively enrolls patients with suspected COVID-19, or COVID-19 confirmed through SARS-CoV-2 PCR testing. For this study, we used results from patients with available WGS data and who were recruited at the Jewish General Hospital (JGH) in Montreal. The JGH is a university affiliated hospital serving a large multi-ethnic adult population and was designated as the primary COVID-19 reference center by the Québec government early in the pandemic. In total, there were 533 participants with WGS, including 62 cases of COVID-19 who required ventilatory support (BiPAP, high flow oxygen, or endotracheal intubation) or died, 128 COVID-19 patients who were hospitalized but did not require invasive ventilatory support, 30 individuals with COVID-19 did not require hospitalization, and 313 SARS-CoV-2 PCR-negative participants. Using genetic principal component analysis derived from genome-wide genotyping, we determined that 76% of participants were of European ancestry, 9% were of African ancestry, 7% were of east Asian ancestry, and 5% were of south Asian ancestry.

We performed WGS at a mean depth of 30x on all included individuals using Illumina’s Novaseq 6000 platform (Illumina, San Diego, CA, USA). Sequencing results were analyzed using the McGill Genome Center bioinformatics pipelines^23^, in accordance with Genome Analysis Toolkit Best Practices (GATK) recommendations^7^. Reads were aligned to the GRCh38 reference genome. Variant quality control was performed using the variantRecalibrator and applyVQSR functions from GATK. Participants had a median of 4434 (IQR: 4260-5194) autosomal single nucleotide polymorphisms of minor allele frequency less than 1% across their exomes (defined using the GENCODE^24^ reference for protein coding exonic sites). Variant filtering was followed by sample filtering on missing rate (minimum 97% of sites called) and minimum average read depth (≥ 20x). Genotypes were further filtered by genotype quality (≥20x) and read depth (≥10x).

Predicted LOF variants were annotated using LOFTEE^10^ and the analysis was restricted to the genes tested in Zhang *et al*. Missense variants were annotated using VEP. Qualifying variants with minor allele frequency < 0.1% annotated as high confidence for LOF or as missense variants and which passed quality control filters were extracted for the main analysis. Using these variants, we used Fisher’s exact test to perform a burden test restricted to individuals of European ancestry comparing 45 of the 62 severe cases of COVID-19 to 361 controls.

### Saudi Arabian COVID-19 Cohort

The Saudi Human Genome Program (SHGP) aims to sequence the genomes of the Saudi patients with COVID-19 confirmed through SARS-CoV-2 PCR as part of COVID19 KACST response (https://covid19.kacst.edu.sa/en.html). The study was conducted in accordance with the ethical principles of the National Bioethical committee at KACST and approved by the Institutional Review Board Committee at King Abdullah International Medical Research Centre, Ministry of National Guard-Health Affairs, Riyadh, Ministry of Health, and King Fahad Medical City. The Institutional Review Boards of all participating hospitals also approved the study and all patients provided written informed consent. In total, we performed WES on 307 individuals with COVID-19, which included 148 (48%) cases of severe disease that required mechanical ventilation or admission to an intensive care unit for septic shock or organ failure and 159 (52%) patients with mild or asymptomatic disease. Our control group included 218 individuals negative by SARS-CoV-2 PCR test. The 159 severe cases included 32% females and 68% males, and the 148 mild/asymptomatic patients were 44% females and 56% males. The age of the 307 patients included in this study ranged from 6 months to 98 years old. All samples were of Arab ancestry.

WES was performed using Ion Torrent S5 XL System (Thermo Fisher Scientific, Carlsbad, CA USA) includes the Torrent Suite™ Software package, which automatically processes sequences, base calling, read alignment, and variant calling. This automated and streamlined data analysis workflow performs annotation and mutation identification on FASTQ, SFF, BAM, and VCF output files. Variant call format (VCF) files were filtered to remove unusually poor-quality or low-coverage reads. All variants retained in the analysis had quality ≥ 30 and coverage ≥ 10. Variant files were annotated using the VEP version 100 and referencing the GRCh37/hg19 build within dbSNP, the NCBI database of genetic variation.

Predicted LOF variants were annotated using LOFTEE^10^ and the analysis was restricted to the 13 genes tested in Zhang et al. Rare variants (minor allele frequency < 1%) annotated as high confidence for LOF by LOFTEE and passed quality control filters were extracted for the main analysis. We scanned for these variants in our entire cohort of control negative, asymptomatic/mild and severe Saudi Arabian COVID-19 patients, and surprisingly, none were present in any patient.

### Qatar Genome Program COVID-19 Cohort

The Qatar Genome Program (QGP) is a population-based project launched by the Qatar Foundation to generate a large whole genome sequence (WGS) dataset, in combination with comprehensive phenotypic information collected by the Qatar Biobank. In this retrospective cohort consisting of 14,060 WGS samples, we extracted information from electronic medical records (EMR) on the participants that were tested positive. The extraction covered the period from early March till early September. There were a total of 700 COVID-19 positive cases, including 60 severe cases with hypoxia that required ventilatory support (BiPAP, high flow oxygen, or endotracheal intubation) and 640 classified as asymptomatic or mild cases without any evidence of pneumonia or hypoxia. The severe COVID-19 cohort consisted of 30 male and 30 female patients that were hospitalized for respiratory failure. Of the mild COVID-19 cases, 290 were male and 350 were female. The median age was 38 (range 18-89) for the severe and 37 (range 18-89) for the mild group. All samples used for the analysis were of Qatari Middle Eastern Arabian ancestry. After removal of related individuals, 23 severe and 231 mild cases remained. The severe COVID-19 cohort consisted of 13 males and 10 females with a median age of 55 (range 28-68). 116 of the mild COVID-19 cohort were male, 115 female with a median age of 37 range (18-79).

All samples were sequenced on Illumina Hiseq X instrument (Illumina, San Diego, CA, USA) to a minimum average coverage of 30x. Quality control of Fastq files was performed using FastQC (v0.11.2). The secondary analyses (read mapping and joint variant calling) were performed using Sentieon’s DNASeq pipeline^25^ v201808.03 (Sentieon, San Jose, CA, USA), following the GATK 3.8 best practices^6,7^. Reads were aligned to GRCh38/hg38 reference genome. Quality control analyses included removal of samples with a low genotyping call rate (less than 95%), gender discordance, excess heterozygosity and PCA-based population outliers. We then used KING^16^ for the detection of related individuals and removed one of each pair that had an inferred relationship of second-degree or closer. Consequently, 5,385 unrelated samples were retained for our analysis. The remaining variants were annotated using VEP (release 99), with LOFTEE and dbNSFP plugins. We then performed a gene set-based collapsing analysis, using the same criteria as described for the Columbia University COVID-19 Biobank Cohort (pLOF with internal & external MAF < 0.1%).

### Ethics statement

The recruitment and sequencing of participants from the Columbia University COVID-19 Biobank were approved by the Columbia University IRB, protocol number AAAS7370. The genetic analyses were approved by the Columbia University IRB, under protocol number AAAS7948. The BQC-19 received ethical approval for its activities from its multicentric ethics board headed at the Centre Hospitalier de l’Université de Montréal. Recruitment of patients at the Jewish General Hospital (JGH) was also approved by the JGH ethics board. The Saudi Arabia COVID-19 Biobank study was conducted in accordance with the ethical principles of the National Bioethical committee at KACST and approved by the Institutional Review Board Committee at King Abdullah International Medical Research Centre, Ministry of National Guard-Health Affairs, Riyadh, Ministry of Health, and King Fahad Medical City. The Institutional Review Boards of all participating hospitals also approved the study. The recruitment and sequencing of participants from the Qatar Biobank (QBB) were approved by the Hamad Medical Corporation Ethics Committee in 2011 and continued with QBB Institutional Review Board (IRB) from 2017 onwards. It is renewed on an annual basis, IRB protocol number IRB-A-QBB-2019-0017. The genetic analyses presented here were approved following expedited review by the QBB Institutional Review Board under protocol number E-2020-QBB-Res-ACC-0226-0130.

## Data Availability

Data are available from the corresponding authors upon reasonable request.

## Acknowledgements

We would like to thank all study participants and their families for contributing to the biobanks used in this study. The sequencing and phenotyping of the Columbia University cohort was made possible by the Columbia University COVID-19 Biobank Genomics Workgroup members, including Andrea Califano, Wendy Chung, Christine K. Garcia, David B. Goldstein, Iuliana Ionita-Laza, Krzysztof Kiryluk, Richard Mayeux, Sheila M. O’Byrne, Danielle Pendrick, Muredach P. Reilly, Soumitra Sengupta, Peter Sims, and Anne-Catrin Uhlemann. We thank the Saudi genome members and collaborators including Sameera Al Johani, Majid Alfadhel, Deema Zahrani, Moneera Alsuwailem, Nouf AlDhawi, Eman Barhoush, Sarah AlKwai, Maha Albeladi, Faisal Almalki, Iman Mohammad, Ahmed Alaskar, Ebtehal A. Alsolm, Sara S. Alotaibi, Aljohara A. Albabtain, Dona Baraka, Rana Hasanato, Hamza A. Alghamdi, Laila S. Al-Awdah, Amin K. Khattab, Roaa Talal Halawani, Ziab Zakey Alahmadey, Jehad Khalid Albakri, Walaa Ali Felemban, and Bandar Ali Suliman. We also thank the Qatar Biobank for data collection and phenotyping. The Qatar Genome Program and Qatar Biobank are both Research, Development & Innovation entities within Qatar Foundation for Education, Science and Community Development. We are also thankful to Dr Laith Abu Raddad for his help in data extraction from EMR.

## Funding

The Columbia University COVID-19 Biobank is supported by the Vagelos College of Physicians & Surgeons Office for Research, Precision Medicine Resource (PMR), and Biomedical Informatics Resource (BMIR) of the Columbia University Irving Institute for Clinical and Translational Research (Columbia CTSA). Columbia CTSA is funded by the National Center for Advancing Translational Sciences, National Institutes of Health, through Grant Number UL1TR001873. A portion of population controls were from the Washington Heights Inwood Columbia Aging Project (AG054023). Whole genome sequencing of the Biobanque Québec Covid-19 cohort was funded by the CanCOGeN HostSeq project, with contribution from the Fonds de Recherche Québec Santé (FRQS), Génome Québec, and the Public Health Agency of Canada. The Richards research group is supported by the Canadian Institutes of Health Research (CIHR), the Lady Davis Institute of the Jewish General Hospital, the Canadian Foundation for Innovation, the NIH Foundation, Cancer Research UK and the FRQS. MA and MSA acknowledge King Abdulaziz City for Science and Technology and the Saudi Human Genome Project for technical and financial support (https://shgp.kacst.edu.sa). MMR was financially supported by King Saud University, Vice Deanship of Research Chairs. GBL is supported by a joint research fellowship from Quebec’s ministry of health and social services, and the FRQS. TN is supported by Research Fellowships of Japan Society for the Promotion of Science (JSPS) for Young Scientists and JSPS Overseas Challenge Program for Young Researchers. JBR is supported by a FRQS Clinical Research Scholarship. This study is part of the www.covid19hg.org consortium.

## Notes

### Competing Interest Statement

The authors have declared no competing interest.

### Summary of Updates

Fixed author name.

